# Evaluating Large Language Models in Interpreting Cervical Cytology

**DOI:** 10.1101/2025.11.04.25339501

**Authors:** Saroja Devi Geetha

## Abstract

**Background:** Large language models (LLMs) have shown promise in medical imaging, but their utility in cytology remains underexplored. This study evaluates GPT-5 and Gemini 2.5 Pro for Pap smear interpretation.

**Methods:** Digital cervical Pap smear images of 100 cases were obtained from the Hologic Education Site, with Hologic diagnoses considered the gold standard. Representative images were uploaded into GPT-5 and Gemini 2.5 Pro and prompted to provide a diagnosis based on the Third Edition of the Bethesda System for Reporting Cervical Cytopathology. Cases with infectious organisms were assessed using additional images. Concordance was evaluated at exact diagnosis and clinical management groupings, wherein diagnoses with similar management implications were grouped. Sensitivity and specificity for abnormal cytology were also calculated.

**Results:** Concordance of both LLMs for exact diagnostic matches were comparable (GPT-5: 47%, Gemini: 48%) and increased to 66% for clinical management grouping. GPT-5 performed best for low-grade squamous intraepithelial lesions (75%), whereas Gemini 2.5 Pro showed the highest concordance in the high-grade squamous intraepithelial lesion (HSIL) category (82%), although this was largely attributable to its strong tendency to overcall cases as HSIL. Sensitivity for detecting abnormal cytology was 74% for GPT-5 and 84% for Gemini, with specificity of 74% and 71%, respectively. GPT-5 better identified glandular lesions, while Gemini detected organisms more accurately (71% vs. 20%).

**Conclusions:** Current LLMs demonstrate moderate ability to identify cytologic abnormalities but are not yet reliable for independent Pap smear interpretation. Targeted fine-tuning, prompt optimization, and cytology-specific training could enhance their utility as adjunctive tools in cytology workflows.

## Introduction

Large language models (LLMs) are advanced artificial intelligence (AI) systems built on transformer architectures, designed to process and generate human language. In recent years, LLMs have gained increasing prominence in the medical domain, demonstrating potential in applications such as generating patient discharge summaries, assisting clinical decision-making, and enhancing medical education.^1,2^

ChatGPT, developed by OpenAI and first introduced in 2018, has undergone multiple revisions. A major advancement came in late 2023 with the release of GPT-4-turbo with vision, which introduced image-analysis capabilities. The current version, GPT-5, launched in August 2025, is trained on a substantially larger and more diverse dataset encompassing specialized domains such as medicine. Along with fully integrated image-understanding mechanisms, GPT-5 represents a truly multimodal LLM, capable of fusing vision, language, and reasoning to support diverse applications in research, diagnostics, education, and data analysis.^3^ Gemini, developed by Google and introduced in late 2023, is another multimodal LLM with built-in visual capabilities.^4^ It includes multiple model variants: Nano, Flash, Pro, and Ultra. The most recent version, Gemini 2.5 Pro, released on March, 2025, combines advanced natural language processing with enhanced analytical performance. ^5^

Pathology, a visually intensive field of medicine, has long been at the forefront of adopting innovations such as whole-slide imaging and AI. Several AI-assisted tools have been successfully implemented in pathology, particularly in cervical cytology, culminating in the recent FDA-approved Hologic Genius Digital Diagnostic System for Pap test screening.^6^ Despite these advances, the potential of LLMs to analyze cervical cytology images remains unexplored. Therefore, this study is aimed to evaluate the ability of GPT-5.0 and Gemini 2.5 Pro in interpreting Pap smear images.

## Methods

Digital images of cervical Pap smears were obtained from the Hologic digital cytology education site. ^7^ A total of 100 cases were included in this study. The Hologic-provided diagnosis for each case was considered the gold standard. For each case, the most representative image (in most cases the ones highlighted for each case by the Hologic site) were selected. The selected images were uploaded into GPT-5^8^ and Google Gemini 2.5 Pro^9^. Each model was prompted to provide a diagnosis based on the Third Edition of the Bethesda System for Reporting Cervical Cytopathology, which includes: Negative for Intraepithelial Lesion or Malignancy (NILM), Atypical Squamous Cells of Undetermined Significance (ASC-US), Low-Grade Squamous Intraepithelial Lesion (LSIL), Atypical Squamous Cells-Cannot Exclude High-Grade Squamous Intraepithelial Lesion (ASC-H), High-Grade Squamous Intraepithelial Lesion (HSIL), Atypical Glandular Cells (AGC), Adenocarcinoma In Situ (AIS), and Adenocarcinoma. The exact prompt used was: “*please analyze this Cervical PAP Smear images and give the diagnosis based on the Third Edition of Bethesda System for Reporting Cervical Cytopathology. Do not include any explanation*.” The diagnoses generated by the LLMs were recorded and compared against the gold standard. For cases demonstrating infectious organisms such as Candida, Trichomonas, and Herpes Simplex Virus (HSV), additional representative images highlighting these organisms were provided to the LLMs for assessment. Concordance of the two LLMs and the gold standard was evaluated at two levels: exact diagnostic agreement, and clinical management level agreement, wherein diagnoses with similar management implications were grouped (ASC-US and LSIL were considered equivalent; ASC-H, HSIL, SCC, AGC, AIS, and adenocarcinoma were grouped together). The ability of each LLM to correctly identify the presence of infectious organisms was also analyzed. Additionally, sensitivity and specificity of each LLM in detecting abnormal cytology was calculated.

## Results

A total of 100 Pap smear cases were included in this study. The distribution of cases across diagnostic categories based on the gold-standard diagnoses are as follows: NILM (n = 43), ASC-US (n = 2), LSIL (n = 20), ASC-H (n = 3), HSIL (n = 22), SCC (n = 4), AGC (n = 3), AIS (n = 1), and adenocarcinoma (n = 3). The diagnoses rendered by GPT-5 were: NILM (n = 35), ASC-US (n= 8), LSIL (n = 25), ASC-H (n = 5), HSIL (n = 20), SCC (n = 0), AGC (n = 6), AIS (n = 0), and adenocarcinoma (n = 1). The diagnoses rendered by Gemini 2.5 Pro were: NILM (n = 25), ASC-US (n = 0), LSIL (n = 23), ASC-H (n = 0), HSIL (n = 51), SCC (n = 0), AGC (n = 0), AIS (n = 0), and adenocarcinoma (n = 3). Table 1 & 2 summarizes the diagnostic distribution and comparison of GPT-5 and Gemini 2.5 Pro against the gold-standard diagnoses respectively. GPT-5 demonstrated an overall concordance rate of 47% (47/100 cases) with the exact gold-standard diagnosis. Gemini 2.5 Pro showed a similar concordance rate of 48% (48/100 cases). Among individual diagnostic categories, GPT-5 achieved the highest concordance in the LSIL category (75%), followed by NILM (61%). In contrast, Gemini 2.5 Pro showed the highest concordance in the HSIL category (82%), although this was largely attributable to its strong tendency to overcall cases as HSIL. The concordance rate for the LSIL category using Gemini 2.5 Pro was 65%.

**Table 1:** Diagnostic distribution of Pap smear cytology interpretations by GPT-5 compared with the gold-standard diagnoses.

| GPT-5<br>Diagnosis | GOLD STANDARD |  |  |  |  |  |  |  |  |
| --- | --- | --- | --- | --- | --- | --- | --- | --- | --- |
|  | NILM<br>(n=43) | ASC-US<br>(N=2) | LSIL<br>(n=20) | ASC-H<br>(n=3) | HSIL<br>(n=22) | SCC<br>(n=4) | AGC<br>(n=3) | AIS<br>(n=1) | ADENO<br>(n=2) |
| NILM (n=35) | 26 | 1 | 1 | 2 | 5 | - | - | - | - |
| ASC-US (n=8) | 4 | - | 2 | - | 2 | - | - | - | - |
| LSIL (n=25) | 5 | - | 15 | 1 | 4 | - | - | - | - |
| ASC-H (n=5) | - | - | - | - | 3 | 2 | - | - | - |
| HSIL (n=20) | 7 | 1 | 2 | - | 5 | 2 | 2 | - | 1 |
| SCC (n=0) | - | - | - | - | - | - | - | - | - |
| AGC (n=6) | 1 | - | - | - | 2 | - | 1 | 1 | 1 |
| AIS (n= 0) | - | - | - | - | - | - | - | - | - |
| AdenoCA (n=1) | - | - | - | - | 1 | - | - | - | - |
| **Concordance** | **61%** | **0%** | **75%** | **0%** | **22%** | **0%** | **33%** | **0%** | **0%** |

Abbreviations- NILM: Negative for Intraepithelial Lesion or Malignancy; ASC-US: Atypical Squamous Cells of Undetermined Significance; LSIL: Low-Grade Squamous Intraepithelial Lesion; ASC-H: Atypical Squamous Cells: Cannot Exclude High-Grade Squamous Intraepithelial Lesion; HSIL: High-Grade Squamous Intraepithelial Lesion; AGC: Atypical Glandular Cells; AIS: Adenocarcinoma In Situ; AdenoCA: Adenocarcinoma.

**Table 2:**
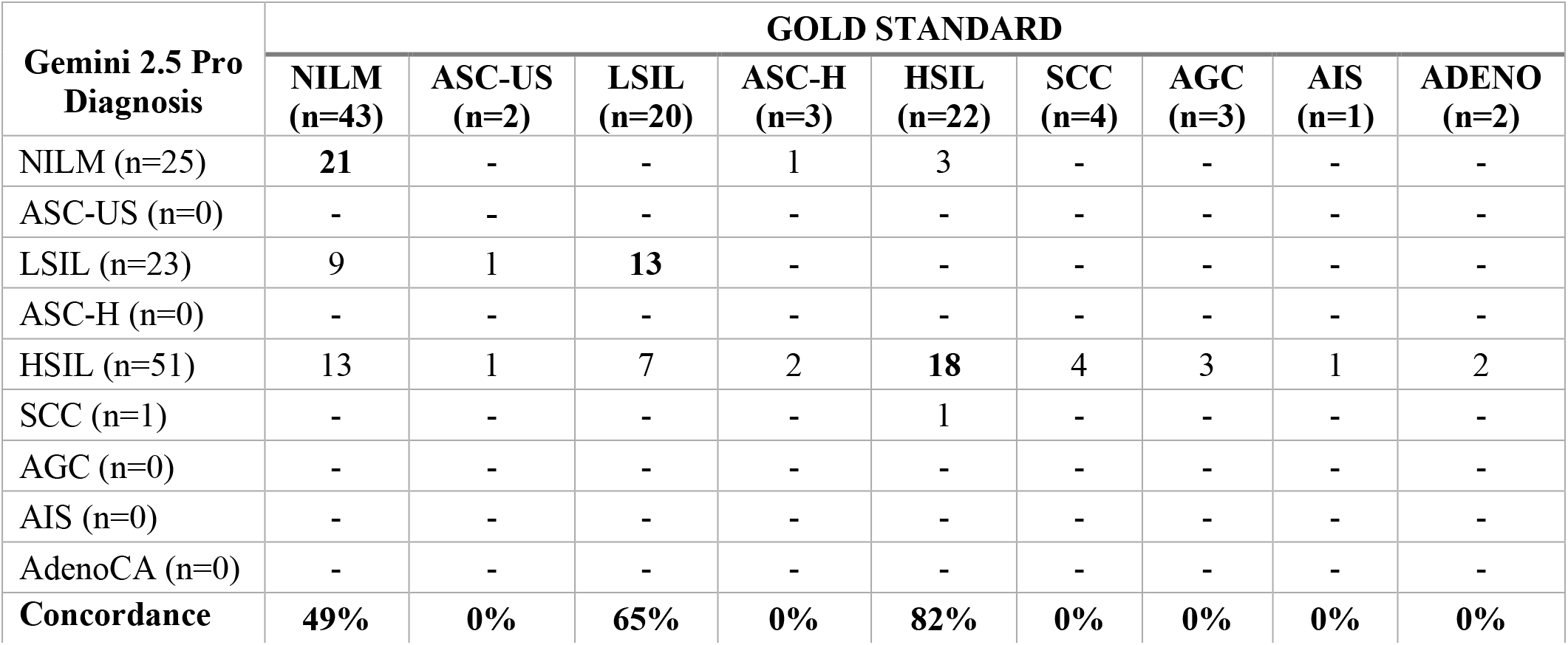
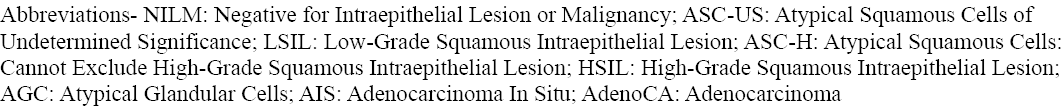
Diagnostic distribution of Pap smear cytology interpretations by Gemini 2.5 Pro compared with the gold-standard diagnoses.

| Gemini 2.5 Pro<br>Diagnosis | GOLD STANDARD |  |  |  |  |  |  |  |  |
| --- | --- | --- | --- | --- | --- | --- | --- | --- | --- |
|  | NILM<br>(n=43) | ASC-US<br>(n=2) | LSIL<br>(n=20) | ASC-H<br>(n=3) | HSIL<br>(n=22) | SCC<br>(n=4) | AGC<br>(n=3) | AIS<br>(n=1) | ADENO<br>(n=2) |
| NILM (n=25) | 21 | - | - | 1 | 3 | - | - | - | - |
| ASC-US (n=0) | - | - | - | - | - | - | - | - | - |
| LSIL (n=23) | 9 | 1 | 13 | - | - | - | - | - | - |
| ASC-H (n=0) | - | - | - | - | - | - | - | - | - |
| HSIL (n=51) | 13 | 1 | 7 | 2 | 18 | 4 | 3 | 1 | 2 |
| SCC (n=1) | - | - | - | - | 1 | - | - | - | - |
| AGC (n=0) | - | - | - | - | - | - | - | - | - |
| AIS (n=0) | - | - | - | - | - | - | - | - | - |
| AdenoCA (n=0) | - | - | - | - | - | - | - | - | - |
| **Concordance** | **49%** | **0%** | **65%** | **0%** | **82%** | **0%** | **0%** | **0%** | **0%** |

Abbreviations- NILM: Negative for Intraepithelial Lesion or Malignancy; ASC-US: Atypical Squamous Cells of Undetermined Significance; LSIL: Low-Grade Squamous Intraepithelial Lesion; ASC-H: Atypical Squamous Cells: Cannot Exclude High-Grade Squamous Intraepithelial Lesion; HSIL: High-Grade Squamous Intraepithelial Lesion; AGC: Atypical Glandular Cells; AIS: Adenocarcinoma In Situ; AdenoCA: Adenocarcinoma

Distribution of cases based on clinical management grouping by GPT-5 and Gemini 2.5 Pro are presented in Table 3 and 4 respectively. When categorized by clinical management relevance, both LLMs demonstrated an overall concordance rate of 66%. Additionally, to assess the LLM’s ability to recognize abnormal cytology, sensitivity and specificity were calculated. The sensitivity of Gemini 2.5 Pro (84%) was slightly higher than that of GPT-5 (74%), while the specificity for both models was comparable (GPT-5: 74%, Gemini 2.5 Pro: 71%).

**Table 3:** Comparison of GPT-5 Pap smear cytology interpretations with gold-standard diagnoses based on clinical management grouping.

| **GPT-5 diagnosis** | **GOLD STANDARD** |  |  |
| --- | --- | --- | --- |
|  | **NILM (n=43)** | **ASCUS/LSIL (n=22)** | **ASC-H and above(n=35)** |
| NILM | **26** | 2 | 7 |
| ASCUS/LSIL | 9 | **17** | 7 |
| ASC-H and above | 8 | 3 | **21** |
| **Concordance** | 61% | 77% | 60% |

Abbreviations- NILM: Negative for Intraepithelial Lesion or Malignancy; ASC-US: Atypical Squamous Cells of Undetermined Significance; LSIL: Low-Grade Squamous Intraepithelial Lesion; ASC-H: Atypical Squamous Cells: Cannot Exclude High-Grade Squamous Intraepithelial Lesion

**Table 4:** Comparison of Gemini 2.5 Pro Pap smear cytology interpretations with gold-standard diagnoses based on clinical management grouping.

| **Gemini 2.5 Pro diagnosis** | **GOLD STANDARD** |  |  |
| --- | --- | --- | --- |
|  | **NILM (n=43)** | **ASCUS/LSIL (n=22)** | **ASC-H and above(n=35)** |
| NILM | **21** | 0 | 4 |
| ASCUS/LSIL | 9 | **14** | 0 |
| ASC-H and above | 13 | 8 | **31** |
| **Concordance** | 49% | 64% | 89% |

Abbreviations- NILM: Negative for Intraepithelial Lesion or Malignancy; ASC-US: Atypical Squamous Cells of Undetermined Significance; LSIL: Low-Grade Squamous Intraepithelial Lesion; ASC-H: Atypical Squamous Cells: Cannot Exclude High-Grade Squamous Intraepithelial Lesion

A total of 14 cases showed the presence of organisms, including Candida (n = 6), HSV (n = 3), Trichomonas (n = 3), and Actinomyces (n = 2). GPT-5 identified organisms in 10 cases, comprising 3 cases each of Candida and Trichomonas; and two cases each of Actinomyces and HSV. However, only 2 of these 10 cases (20%) were concordant with the gold-standard diagnosis: one Candida and one HSV (Table 5). Among the remaining 8 discordant cases, 6 showed no organisms on the gold standard, and in 2 cases the organisms were misclassified (Actinomyces identified as Candida and Candida identified as Actinomyces). Gemini 2.5 Pro reported organisms in 7 cases, including Candida (n = 6) and Trichomonas (n = 1). Of these, 5 cases (71%) were concordant with the gold-standard diagnosis (one Trichomonas and four Candida). In the remaining two cases, Gemini 2.5 Pro identified Candida where no organisms were present on the gold standard diagnosis (Table 6).

**Table 5:** Detection of infectious organisms by GPT-5 compared with gold-standard diagnoses.

| **GPT-5 interpretation** | **GOLD STANDARD** |  |  |  |
| --- | --- | --- | --- | --- |
|  | **Candida (n=6)** | **Trichomonas (n=3)** | **HSV (n=3)** | **Actinomyces (n=2)** |
| Candida | 1 | - | - | 1 |
| Trichomonas | - | - | - | - |
| HSV | - | - | 1 | - |
| Actinomyces | 1 | - | - | - |
| No organism | 4 | 3 | 2 | 1 |
| **Concordance** | 17% | 0% | 33% | 0% |

Abbreviations- HSV: herpes simplex virus

**Table 6:** Detection of infectious organisms by Gemini 2.5 Pro compared with gold-standard diagnoses.

| Gemini 2.5 Pro interpretation | GOLD STANDARD |  |  |  |
| --- | --- | --- | --- | --- |
|  | Candida (n=6) | Trichomonas (n=3) | HSV (n=3) | Actinomyces (n=2) |
| Candida | 4 | - | - | - |
| Trichomonas | - | 1 | - | - |
| No organism | 2 | 2 | 3 | 2 |
| Concordance | 67% | 33% | 0% | 0% |

Abbreviations- HSV: herpes simplex virus

## Discussion

This study demonstrates that, in their current form, LLMs such as GPT-5 and Gemini 2.5 Pro are not yet suitable for independent interpretation of cytology Pap smears. Both models showed comparable overall performance; however, their diagnostic tendencies differed. Gemini 2.5 Pro displayed a higher sensitivity (84%) than GPT-5 (74%), suggesting a greater ability to detect abnormal cytology. Cervical cytology, unlike histopathology, is inherently prone to significant inter-observer and intra-observer variability, particularly in borderline categories such as ASC-US vs. LSIL, ASC-H vs. HSIL or HSIL vs AGC.^10^ To address this, diagnostic categories were consolidated based on clinical management relevance, which improved the models’ concordance rates from 44% to 66%. This finding highlights the potential of these LLMs for further fine-tuning and training to enhance diagnostic performance in the future.

Gemini 2.5 Pro was limited to providing diagnoses within only four categories: NILM, LSIL, HSIL, and SCC and did not assign diagnoses outside these categories. Notably, Gemini exhibited a strong bias towards HSIL, classifying 51% of cases as such. While this led to a high concordance for HSIL (missing only three cases), it also resulted in numerous false positives, including misclassifying 13 NILM cases as HSIL. The primary reasons for these misclassifications were the presence of atrophy (5/13) and viral cytopathic changes due to HSV (2/13), both of which can mimic HSIL features for the untrained^11^ (Figure 1). In contrast, GPT-5 classified only 5 of these 13 NILM cases as HSIL (two with atrophy and one with HSV), suggesting relatively better contextual interpretation in such challenging scenarios.

**Figure 1:**
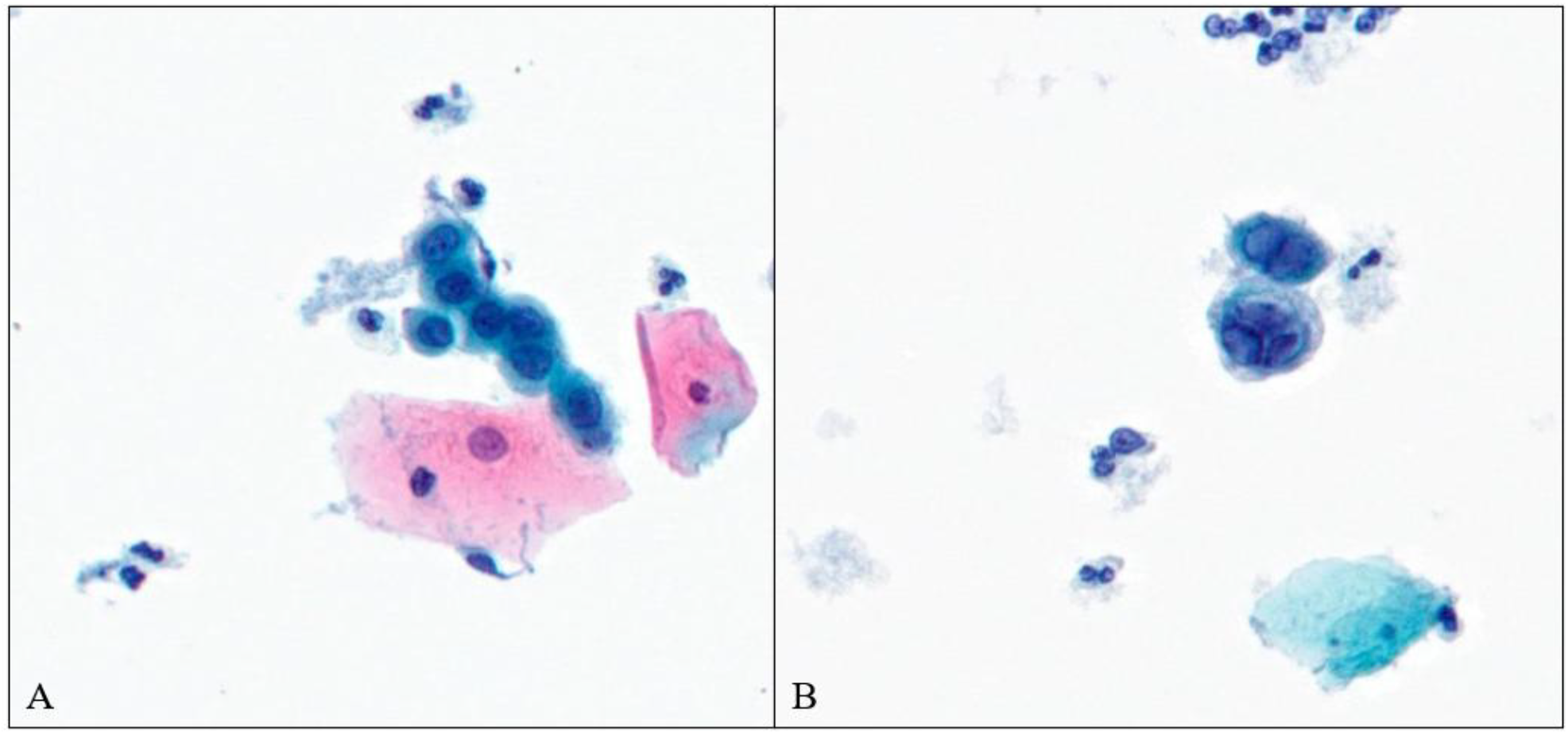
Cases of negative for intraepithelial lesion or malignancy misclassified as high-grade squamous intraepithelial lesion by Gemini 2.5 Pro. (A) A case with atrophy (B) A case showing classic herpetic cytopathic changes

GPT-5 was able to classify smears across a wider spectrum of Bethesda diagnostic categories compared to Gemini 2.5 Pro. Its highest concordance was seen with LSIL (75%) and outperformed Gemini’s LSIL concordance (65%). However, GPT-5’s performance was weakest for HSIL (22% concordance), with several false negatives including five HSIL cases misclassified as NILM, an error with serious clinical implications. For NILM, GPT-5 achieved a moderate concordance of 61%, though it also misclassified seven NILM cases as HSIL, further underscoring the model’s limitations in differentiating high-grade lesions from benign mimics.

Regarding glandular lesions, GPT-5 outperformed Gemini. GPT-5 correctly classified 3 of 6 cases under the AGC category and the remaining 3 as HSIL, whereas Gemini classified all 6 as HSIL. Distinguishing glandular from high-grade squamous lesions remains a well-recognized challenge even for experienced cytopathologists^12^. Nevertheless, when classic glandular features such as nuclear elongation and feathering are present, a diagnosis of AIS is generally straightforward. In this study, a case of AIS with these classic features was identified by GPT-5 as AGC but misclassified as HSIL by Gemini (Figure 2).

**Figure 2:**
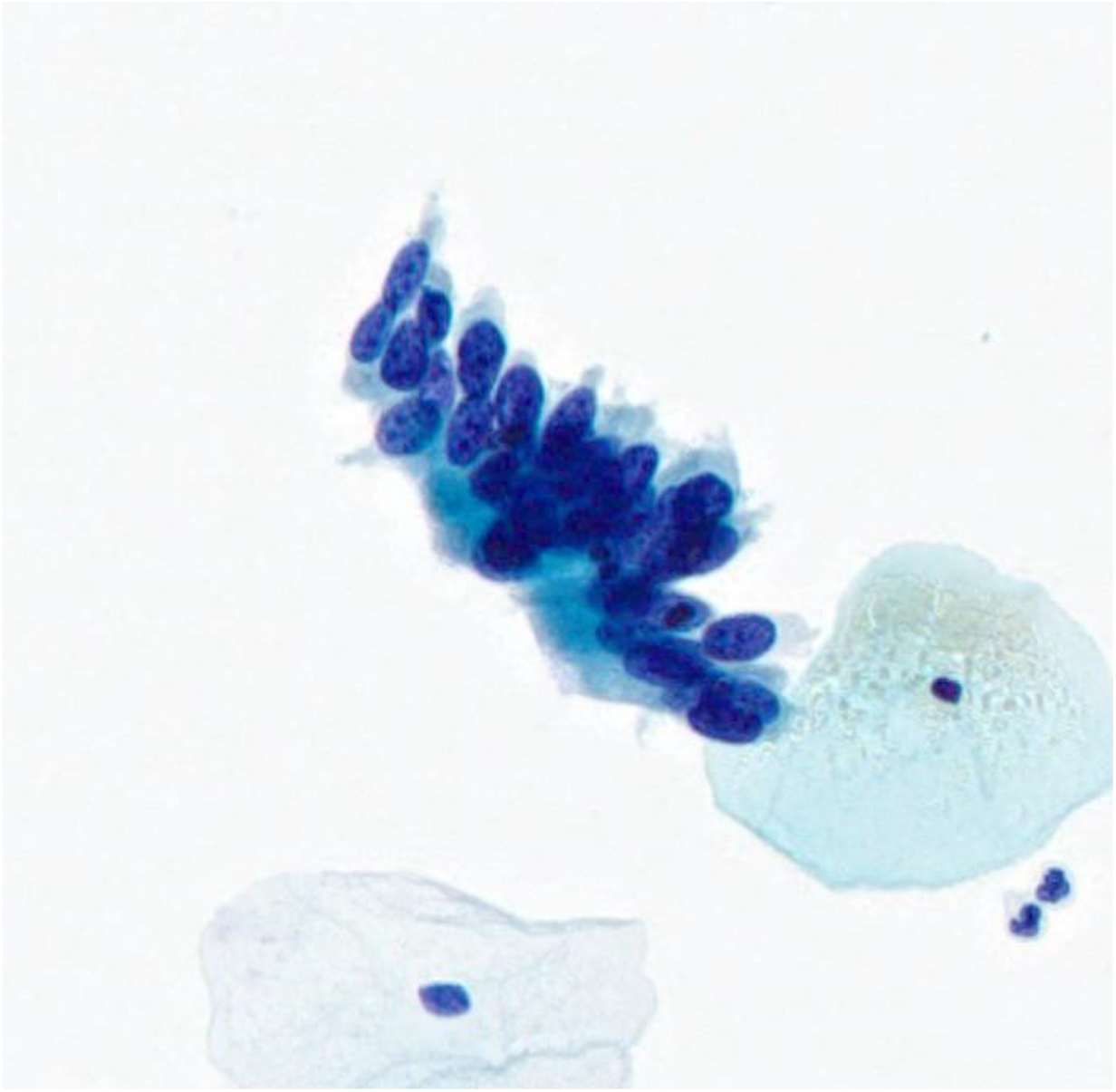
Image from a case of adenocarcinoma in situ. GPT-5 interpreted the case as atypical glandular cells, whereas Gemini 2.5 Pro classified it as high-grade squamous intraepithelial lesion.

Finally, in the detection of infectious organisms, Gemini 2.5 Pro demonstrated superior accuracy (71%) compared to GPT-5 (20%). GPT-5 showed a tendency to both overlook organisms and to interpret their presence when absent, suggesting a need for refinement in image feature recognition related to microorganisms.

While LLMs have shown promise in natural language processing tasks in medicine, there are very few studies that specifically examine their capabilities in pathology image recognition, highlighting a major gap in literature. To the best of knowledge, only one prior study has evaluated the role of LLMs in Pap smear cytology, specifically assessing GPT-3 and Gemini 2.5 Pro. ^13^ Although their overall findings aligns with this study in demonstrating the limited performance of current LLMs, the category-wise results differ. In that study, GPT’s highest accuracy was reported for NILM (93%), while performance for LSIL was markedly lower (26%), which contrasts with the findings of this study where LSIL accuracy was the highest. These discrepancies are likely attributable to differences in the GPT versions tested and model training architectures. They also reported challenges in interpreting glandular lesions, a limitation consistent with the observations of this study.

Limited studies have explored LLM capabilities in histopathology image interpretation.^14^ One such study evaluated GPT-4 for identifying tissue origin and classifying colorectal biopsies, achieving an overall accuracy of 64%, with classification accuracy for polyp subtyping ranging from 57-75%, performance comparable to pathology residents.^15^ Similarly, the application of LLMs in hematopathology has shown promise; demonstrating that GPT-4 can assist not only in recognizing abnormal blood morphology with high accuracy but also in providing relevant diagnostic context regarding possible etiologies and clinical correlates.^16^

In addition to fine tuning and training the LLM models, choosing the correct prompts defined as “prompt engineering” has been known to have influence on their performance.^17,18^ This holds true for pathology image interpretation as well. A study evaluating two different prompts for identifying hepatobiliary tumors using identical image sets found that diagnostic responses varied considerably with prompt structure. Incorporating morphologic descriptive cues into prompts significantly enhanced GPT-4’s diagnostic accuracy.^19^ Another recent study assessed GPT-4o and Claude 3.5 Sonnet for interpreting thyroid fine-needle aspiration cytology, demonstrating improved diagnostic performance following prompt optimization. ^**20**^

Together, these findings highlight that while LLMs currently exhibit variable accuracy across cytology and histopathology applications, their diagnostic potential can be substantially improved through targeted fine-tuning and prompt refinement. Our study adds to this growing body of evidence by emphasizing the unique challenges of cytology interpretation, particularly the morphological challenges and diagnostic gray zones that demand contextual understanding. Given the limited number of studies investigating LLM based image recognition in cytology, these findings provide important insights and underscore the need for future research integrating domain-specific datasets and optimized prompt frameworks to enhance the utility of LLMs in cytopathology practice.

## Conclusion

In conclusion, this study demonstrates that current LLMs, including GPT-5 and Gemini 2.5 Pro, are not yet reliable for independent interpretation of Pap smear cytology. While both models showed moderate concordance with human diagnoses when diagnostic categories were consolidated, significant limitations remain, particularly in identifying high-grade lesions and glandular abnormalities, which carry critical clinical implications. GPT-5 performed better in distinguishing LSIL and glandular lesions, whereas Gemini showed higher accuracy in organism detection but overcalled HSIL. These findings, together with the limited number of studies on LLM-based image recognition in pathology, highlight both the current challenges and future potential of LLMs in cytopathology. With domain-specific fine-tuning, optimized prompt engineering, and integration of curated datasets, LLMs may become a valuable adjunct in cytology workflows, supporting pathologists in screening and diagnostic decision-making.

## Data Availability

All data produced in the present study are available upon reasonable request to the authors

## References

1. Shool S, Adimi S, Saboori Amleshi R, Bitaraf E, Golpira R, Tara M. A systematic review of large language model (LLM) evaluations in clinical medicine. BMC Med Inform Decis Mak. Mar 07 2025;25(1):117. doi:10.1186/s12911-025-02954-4

2. Geetha SD, Khan A, Kannadath BS, Vitkovski T. Evaluation of ChatGPT pathology knowledge using board-style questions. Am J Clin Pathol. Apr 03 2024;161(4):393–398. doi:10.1093/ajcp/aqad158

3. Wang S, Hu M, Li Q, Safari M, Yang X. Capabilities of gpt-5 on multimodal medical reasoning. arXiv preprint arXiv:250808224. 2025;

4. Perera P. Preparing to revolutionize education with the multi-model GenAI tool Google Gemini? A journey towards effective policy making. 2023;

5. Team G, Anil R, Borgeaud S, et al. Gemini: a family of highly capable multimodal models. arXiv preprint arXiv:231211805. 2023;

6. Cantley RL, Jing X, Smola B, Hao W, Harrington S, Pantanowitz L. Validation of AI-assisted ThinPrep® Pap test screening using the Genius. J Pathol Inform. Dec 2024;15:100391. doi:10.1016/j.jpi.2024.100391

7. Hologic Digital Cytology Education Site. Accessed Oct 26 2025. https://www.digitalcytologyeducation.com/

8. ChatGPT-5 OA. Accessed Oct 26, 2025. https://chatgpt.com/?oai-dm=1

9. Google Gemini 2.5 Pro. Accessed Oct 26, 2025. https://gemini.google.com/app

10. Deepthi V, Ramaswamy A, Prashant B. Inter observer variability in the interpretation of atypical squamous cells in Pap smear examination. 2013;

11. Torous VF, Pitman MB. Interpretation pitfalls and malignant mimics in cervical cytology. J Am Soc Cytopathol. 2021;10(2):115–127. doi:10.1016/j.jasc.2020.06.005

12. Bansal B, Gupta P, Gupta N, Rajwanshi A, Suri V. Detecting uterine glandular lesions: Role of cervical cytology. Cytojournal. 2016;13:3. doi:10.4103/1742-6413.177156

13. Laohawetwanit T, Apornvirat S, Asaturova A, Li H, Lami K, Bychkov A. Evaluation of general-purpose large language models as diagnostic support tools in cervical cytology. Pathol Res Pract. Oct 2025;274:156159. doi:10.1016/j.prp.2025.156159

14. Laohawetwanit T, Namboonlue C, Apornvirat S. Accuracy of GPT-4 in histopathological image detection and classification of colorectal adenomas. J Clin Pathol. Feb 18 2025;78(3):202–207. doi:10.1136/jcp-2023-209304

15. Ding L, Fan L, Shen M, et al. Evaluating ChatGPT’s diagnostic potential for pathology images. Front Med (Lausanne). 2024;11:1507203. doi:10.3389/fmed.2024.1507203

16. Yang WH, Yang YJ, Chen TJ. ChatGPT’s innovative application in blood morphology recognition. J Chin Med Assoc. Apr 01 2024;87(4):428–433. doi:10.1097/JCMA.0000000000001071

17. Russe MF, Reisert M, Bamberg F, Rau A. Improving the use of LLMs in radiology through prompt engineering: from precision prompts to zero-shot learning. Rofo. Nov 2024;196(11):1166–1170. doi:10.1055/a-2264-5631

18. Nguyen D, MacKenzie A, Kim YH. Encouragement vs. liability: How prompt engineering influences ChatGPT-4’s radiology exam performance. Clin Imaging. Nov 2024;115:110276. doi:10.1016/j.clinimag.2024.110276

19. Laohawetwanit T, Apornvirat S, Namboonlue C. Thinking like a pathologist: Morphologic approach to hepatobiliary tumors by ChatGPT. Am J Clin Pathol. Jan 28 2025;163(1):3–11. doi:10.1093/ajcp/aqae087

20. Dalal BS, Mukhopadhyay K, Roy D, Bhattacharya S, Chakrabarti I, Mondal SK. Prompt engineering and diagnostic accuracy of multimodal large language models in thyroid fine-needle aspiration cytology. Bioinformation. 2025;21(6):1317–1323. doi:10.6026/97320630021317

